# Beneficial influence of single-stage posterior surgery for the treatment of lumbar brucella spondylitis combined with spondylolisthesis

**DOI:** 10.1101/2022.05.12.22274999

**Authors:** Yao Zhang, Chang-song Zhao, Jia-min Chen, Qiang Zhang

## Abstract

We aimed to evaluate the clinical efficacy of the single-stage posterior surgical treatment for patients of lumbar brucella spondylitis combined with spondylolisthesis. In this study, we performed a retrospective analysis of 16 patients with lumbar brucellosis spondylitis combined with spondylolisthesis from January 2015 to January 2019. All patients underwent single-stage posterior lumbar debridement, reduction, interbody fusion, and instrumentation. Preoperative and postoperative of the visual analog scale (VAS), the Japanese orthopedic association scores (JOA), the Oswestry disability index (ODI), erythrocyte sedimentation rate (ESR), and C-reactive protein (CRP) were compared. In addition, the spondylolisthesis reduction rate, reduction loss rate, interbody fusion rate, and complication rate were recorded. VAS, JOA, ODI, ESR, and CRP were conducted with repeated analysis of variance data at different follow-ups. The postoperative follow-up was 12 to 36 months, with an average of (25.0±8.1) months. VAS, JOA, ODI, ESR, and CRP were significantly better at two-week and one-year follow-up than preoperative results (*P*=0.000, respectively). In addition, one year after the operation, VAS, JOA, ODI, ESR, and CRP showed a significant improvement (*P*=0.000, respectively). The average spondylolisthesis reduction in two weeks after operation was (91.2±6.7) %, and the median reduction loss rate in one year after operation was 8.0 (5.0,9.8) %. At the last follow-up, all patients achieved interbody fusion, no loosening and fracture of instrumentation were found, and no recurrence happened. Single-stage posterior operation for lumbar debridement, reduction, interbody fusion, and instrumentation is beneficial for treating lumbar brucellosis spondylitis combined with spondylolisthesis. Furthermore, the reconstruction of spinal stability may relieve pain, heal lesions, and improve patients’ living.

## Introduction

Brucellosis is a common zoonosis affecting half a million people annually. The disease is transmitted to humans through direct/indirect contact with infected animals or raw meat and dairy product consumption. In China, the brucellosis epidemic mainly exists in Inner Mongolia Autonomous Region and Hebei Province. Brucellosis could enter the human body through the respiratory tract, skin, and digestive tract, resulting in human fever and multiple organ damage [1,2]. Brucella mainly invades large joints, and the most frequently affected part is the spine [3]. The incidence of brucella spondylitis is 2% ∼ 60% [4,5], which is prone to the lumbar spine. Due to the apparent destruction of the upper and lower edges of the vertebral body, it is easy to cause spinal instability [6].

At present, antibiotic treatment combined with surgical treatment is still the primary method for treating Brucella spondylitis, and the clinical efficacy is not satisfactory for patients with antibiotic treatment only. The patients with brucella spondylitis require surgical treatment, accounting for 3% ∼ 29% [4]. After spinal infection with Brucella, necrotic tissue or intervertebral disc destruction occurs in the spinal canal. These lesions will cause indirect or direct compression of the nerve and neurological symptoms. In addition, antibiotic treatment can not relieve the symptoms caused by the compression of the spinal cord, cauda equina nerve, and nerve root, so surgical treatment is required [7].

Lumbar spondylolisthesis is one of the common causes of low back and leg pain, and the specific pathogenesis is unclear. Its classification and treatment methods are diverse. The treatment principle is to restore the spinal sequence as much as possible and eliminate the causes of pain [8]. At present, there is no unified theory about the choice of conservative and surgical treatment, the choice of surgical approach, the choice of minimally invasive and open, and the choice of fusion and decompression [9].

So far, there are few reports on surgical treatment of lumbar brucella spondylitis combined with spondylolisthesis. This study evaluated the clinical efficacy of the single-stage posterior surgical treatment for lumbar brucella spondylitis combined with spondylolisthesis.

## Materials and methods

### Patient population

From January 2015 to January 2019, we included 212 patients with lumbar brucella spondylitis in our department. 142 patients were under antibiotic treatment, and 70 were treated with surgery. Patients met the following indications for surgery: 1. Persisting low back pain; 2. Progressive neurological deficit; 3. Spinal instability; 4. Paravertebral abscesses not easily absorbed; 5. Poor outcomes following antibiotic treatment (ESR and CRP not significantly decreased). 16 cases of lumbar brucella spondylitis combined with spondylolisthesis underwent single-stage posterior debridement, reduction, interbody fusion, and instrumentation. (**Fig 1**)

**Figure 1.**
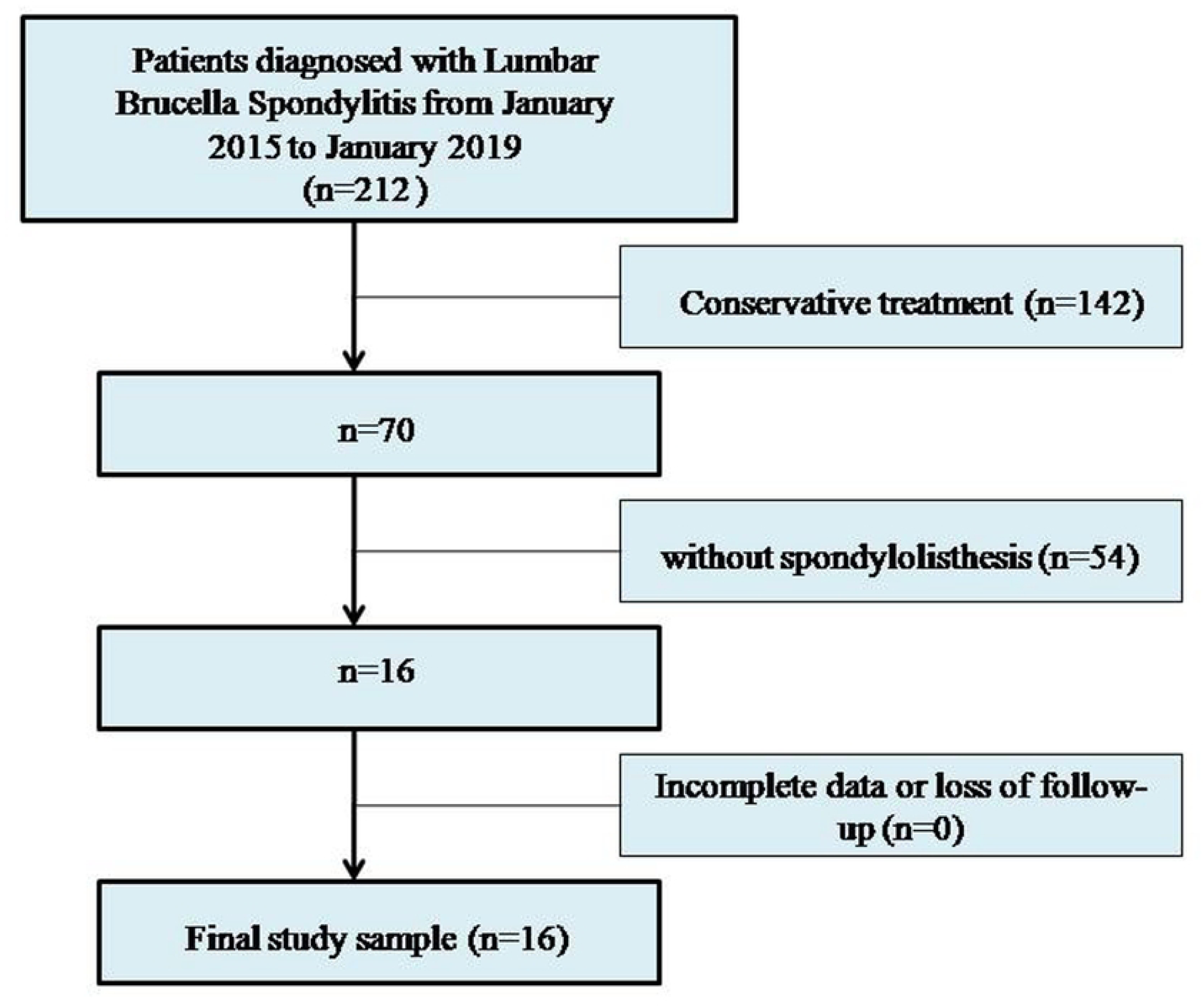
Flowchart showing study enrollment.

The inclusion criteria were as follows: 1. Epidemic history: living in or having been to pastoral areas, having exposure to cattle and sheep, or eating non sterilized cattle, mutton, or dairy products; 2. Symptoms and signs: fever, sweats, fatigue, weight loss, low back pain, and lower limb nerve symptoms; 3. Imaging findings: preoperative X-ray, CT, and MRI findings were consistent with the signs of brucella spondylitis and combined with spondylolisthesis; 4. Serological and microbiological evidence: Rose Bengal plate agglutination test (RBP) was positive or histopathological examination was positive; 5. No other etiological findings (such as Mycobacterium tuberculosis, Staphylococcus aureus, fungi, et al.). (**Table 1**)

**Table 1.**
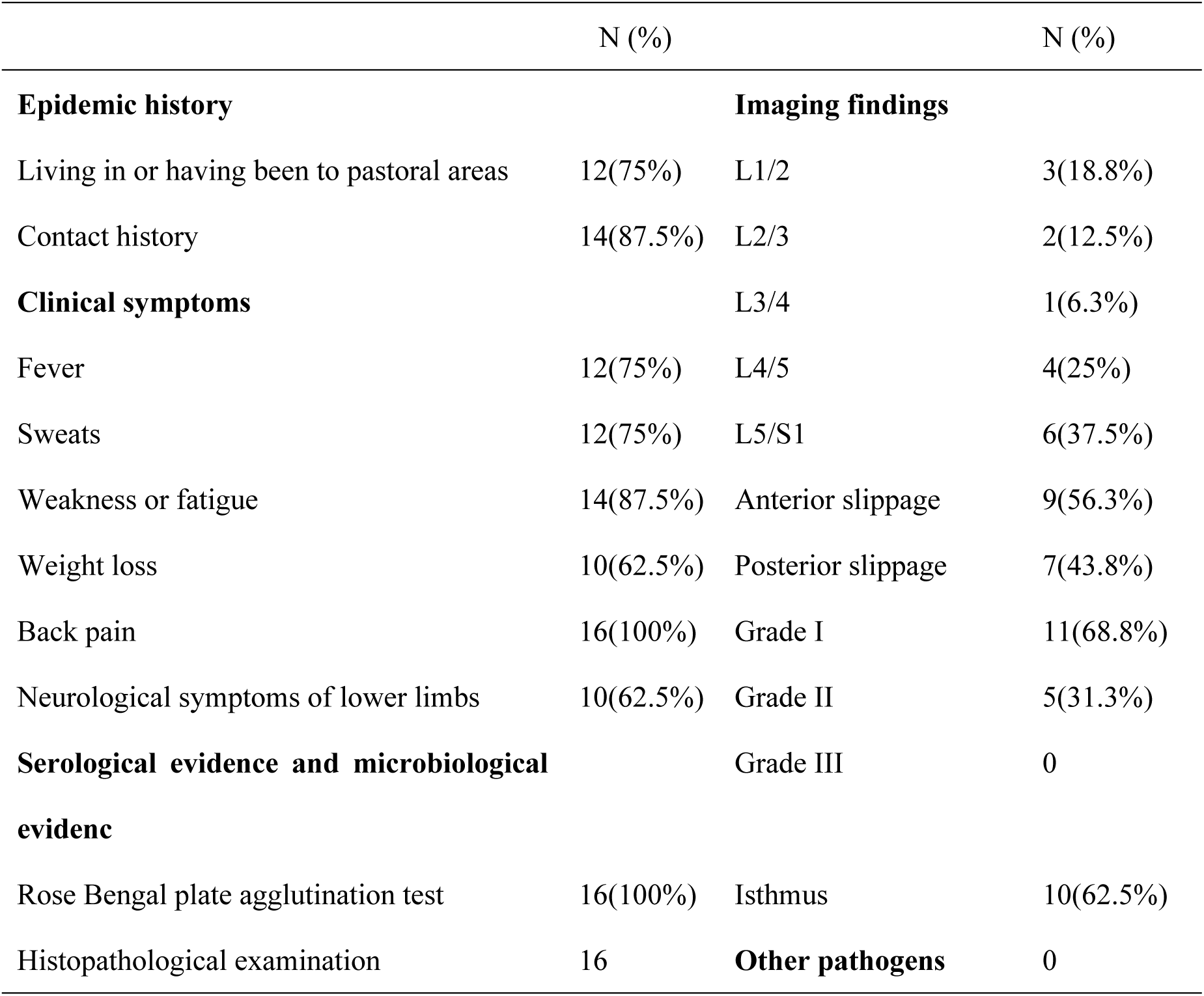
Clinical data of patients.

All patients in this study voluntarily signed a written informed consent form to join the scientific research and signed a written informed consent form for surgical treatment.

### Data collection

The operation time, intraoperative bleeding, and complications were recorded. The patients were followed up regularly at 2 weeks, 1 month, 2 months, 3 months, 6 months, and 1 year after the operation. During the follow-up, X-rays of the lumbar spine were performed in the frontal, lateral and dynamic positions, and a CT scan of the lumbar spine would be performed if necessary. We calculated the spondylolisthesis reduction rate 2 weeks after operation by measuring the vertebral body slippage, i.e. (preoperative slippage - postoperative slippage) / preoperative slippage × 100% [10]. The spondylolisthesis rate at 2 weeks and 1 year after operation was measured by Taillard’s measurement [11], and the reduction loss rate was also calculated. In addition, we observed the condition of the fusion of bone graft and checked the instrumentation 2 weeks and 1 year after the operation [12,13]. The visual analysis scale (VAS), the Japanese Orthopaedic Association scores (JOA), the Oswestry disability index (ODI), and laboratory tests included preoperative and postoperative erythrocyte sedimentation rate (ESR), and C-reactive protein (CRP) were recorded.

### Preoperative preparation

Patients were treated with combinations of antibiotics. The regimens included oral doxycycline 200 mg/day, oral rifampicin 600mg/day, and intravenous levofloxacin 500mg/day, or plus intravenous ceftriaxone sodium 2g/day for at least 6 weeks. In addition, nutrition enhancement and correction of anemia and hypoproteinemia in needed patients were also carried out.

### Surgical techniques

Under general anesthesia, the patients were placed in the prone position. The pedicle of the spondylolisthesis vertebral body was determined and marked fluoroscopically. First, the posterior median incision was made. Then, the skin, subcutaneous, and fascia layers were incised successively and separated to both sides along the spinous process until the lamina and facet joints. Then reduction screws were placed on both sides of the spondylolisthesis vertebral body, and ordinary pedicle screws were placed in the adjacent vertebral body. Subsequently, unilateral or bilateral fenestration decompression was performed. Then, the inflammatory tissues in the spinal canal, intervertebral space, paravertebral and damaged bone were removed entirely, and the intervertebral space was removed until there was blood exudation. Then, we repeatedly washed it with a flushing gun, dried it, installed the longitudinal connecting rod, and lifted and restored the spondylolisthesis vertebral body while opening the intervertebral space properly. Finally, the bitten autologous bone was implanted into the bone defect between vertebrae, and then the polyetheretherketone (PEEK) cage was obliquely inserted. After moderate compression, it was tightened and fixed. Then, we rewashed the gun. After checking that there was no active bleeding, we placed a drainage tube routinely and sutured the incision layer by layer. In the end, the tissues taken during the operation were sent for pathological examination. (**Fig.2**)

**Figure 2.**
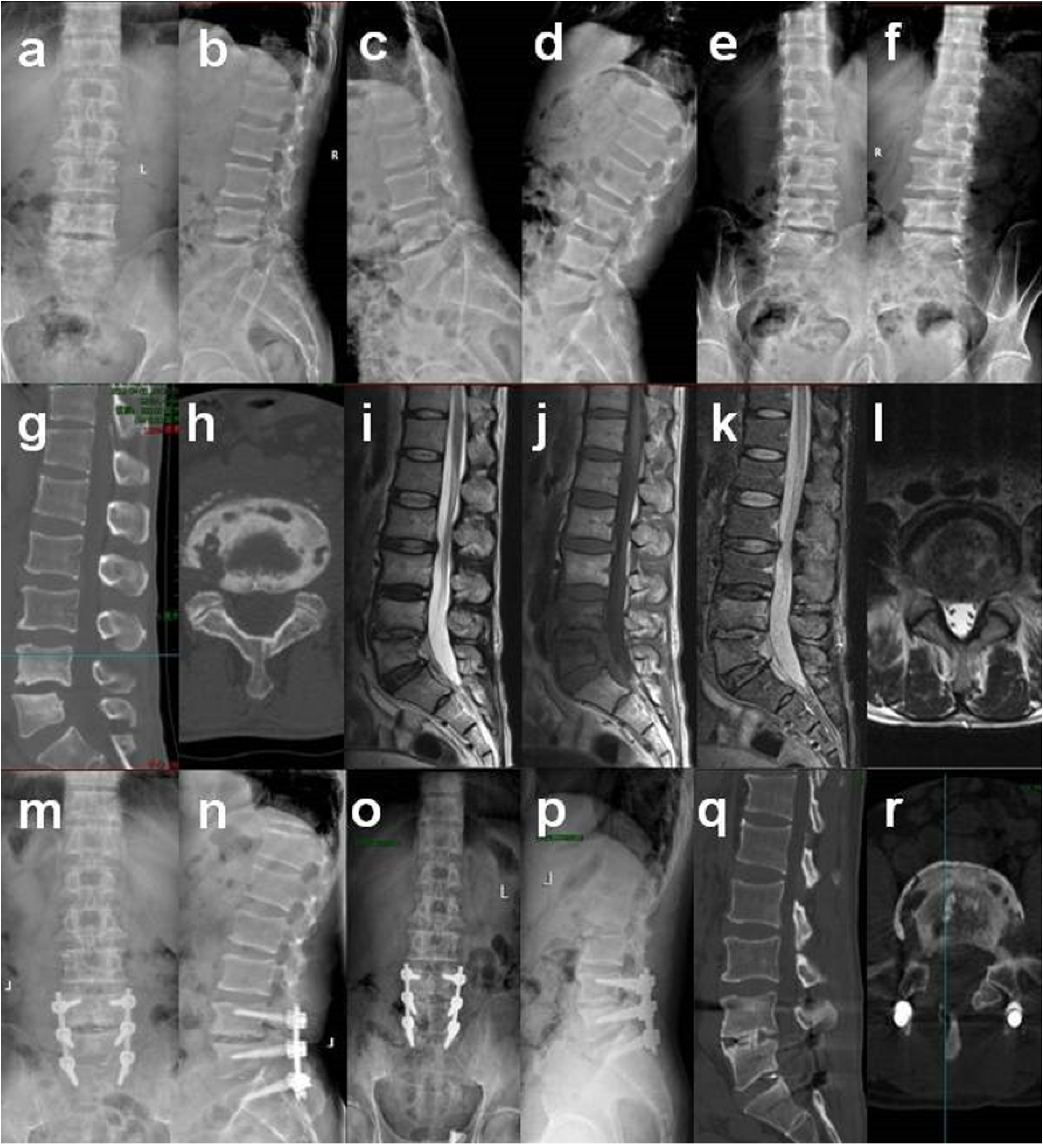
Preoperative and postoperative imaging findings of typical patients. **a-f** Preoperative X-ray showed L4 vertebral body slipped backward II, lumbar instability in a dynamic position, and no obvious isthmus was found in a double oblique position. **g-h** Preoperative CT showed multiple lesions at the upper and lower edges of the vertebral body. **i-l** Preoperative MRI showed uneven abnormal signal changes in the vertebral body and intervertebral disc. **m-n** Two weeks after the operation, X-ray showed L4 spondylolisthesis reduction. **o-p** One year after the operation, X-ray showed there was no apparent loss of spondylolisthesis reduction, and the spine was stable. **q-r** One year after the operation, CT showed interbody fusion had been achieved, and the damaged area was repaired well.

### Statistical analysis

All statistical analyses were performed using the SPSS software (version 25.0; IBM SPSS, New York). Continuous variables were presented as mean ± standard deviation or medians with interquartile ranges, while categorical variables as the frequencies or percentages of events. Mann-Whitney U test was used for nonnormally distributed continuous variables and a t-test for normally distributed variables. VAS, JOA, ODI, ESR, and CRP were conducted with repeated measures analysis of variance data. The *P* value ≤ 0.05 was considered to indicate statistical significance.

## Results

### Basic information

Among the 16 patients, there were 14 males and 2 females. The age ranged from 46 to 68 years, with an average of (59.2 ± 6.5) years. The follow-up time of 16 patients ranged from 12 to 36 months, with an average of (25.0 ± 8.1) months. In addition, 12 patients had a history of living in or having been to pastoral areas, and 14 patients had a history of having exposure to cattle and sheep or eating non sterilized cattle, mutton, or dairy products.

All patients had back pain (100%), followed by weakness or fatigue (87.5%), fever (75%), sweats (75%), then weight loss and neurological symptoms of lower limbs (62.5%, respectively).

There were 3 cases of L1 / 2 slippage, 2 cases of L2 / 3 slippage, 1 case of L3 / 4 slippage, 4 cases of L4 / 5 slippage, and 6 cases of L5 / S1 slippage. There were 9 cases of anterior slippage and 7 cases of posterior slippage, including 10 cases with the isthmus. In addition, there were 11 cases with grade I, 5 with grade II, and no grade III, according to Meyerding grade [14]. (**Table 1**)

### Surgical data and findings

The operation time of 16 patients ranged from 1.5 h to 3.0 h, the median was 2.0 (1.5,2.0) h, the amount of bleeding was 300 ml ∼ 550 ml, and the average was (403.1 ± 64.2) ml. The incisions of 14 patients healed in the first stage, and 2 patients healed delayed due to local scabs. There were no related complications, and no spinal cord, cauda equina, nerve root, and vascular injury were found early after the operation.

The pathological HE staining of 16 patients showed many inflammatory cells such as monocytes, lymphocytes, and neutrophils in the lesion area, which was consistent with the changes in brucellosis. Brucella could be seen by Giemsa staining. (**Fig.3**)

**Figure 3.**
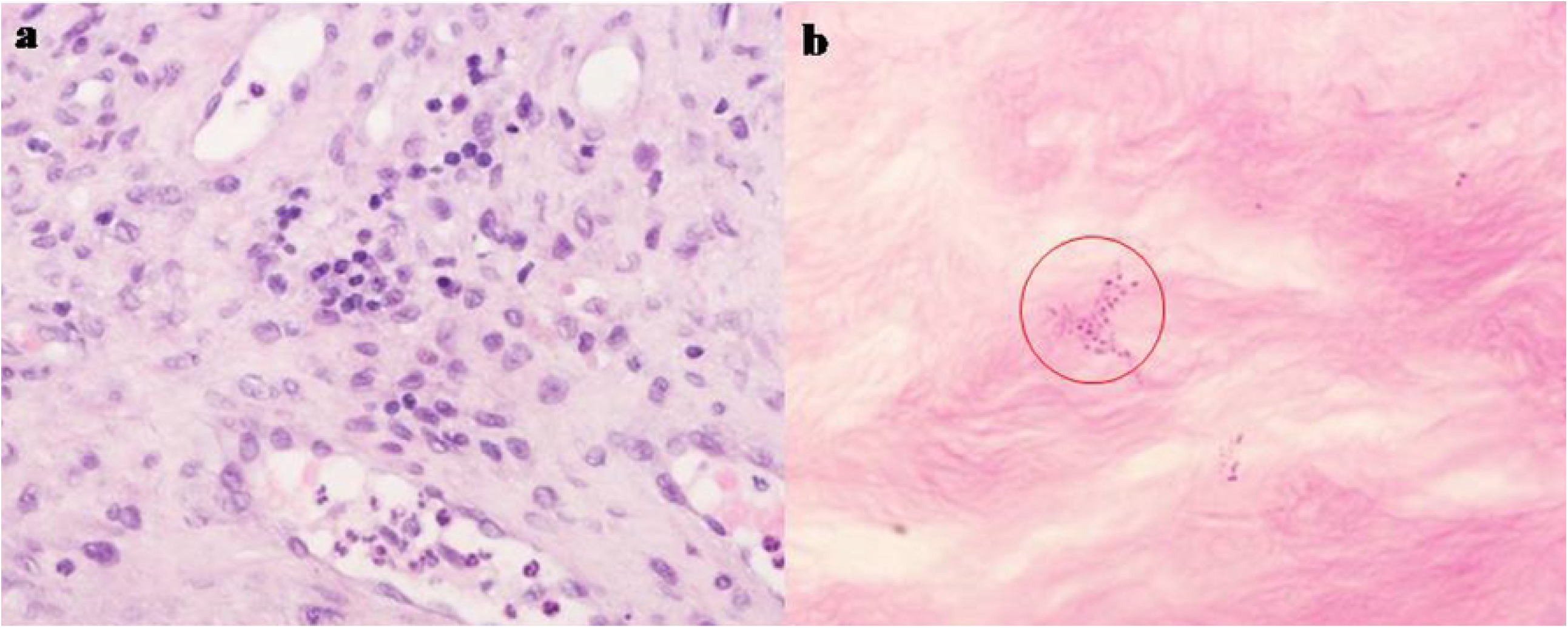
Histopathological results of postoperative lesions. **a** HE staining showed many different types of inflammatory cells in the lesions (x400). **b** Brucella saw by Giemsa staining (x1000)

### Comparison of preoperative and postoperative lumbar function index

The VAS score decreased from 8.0 (8.0,8.8) preoperatively to 2.0 (1.3,2.0) and 0.0 (0.0,1.0) at 2 weeks and 1 year after the operation, respectively. The JOA score increased from the average (11.8 ± 3.6) preoperatively to (18.6 ± 2.3) and (23.6 ± 2.7) at 2 weeks and 1 year after the operation, respectively. ODI index decreased from (88.5 ± 5.6) % preoperatively to (35.7 ± 3.1) % and (9.3 ± 5.7) % at 2 weeks and 1 year after the operation, respectively. VAS score, JOA score, and ODI index at 2 weeks and 1 year after the operation were significantly different from those before the operation (*P* = 0.000) (**Table 2**). Moreover, 1 year after the operation, the VAS score, JOA score, and ODI index were significantly different from those 2 weeks after the operation (*P* = 0.000) (**Table 2**).

**Table 2.**
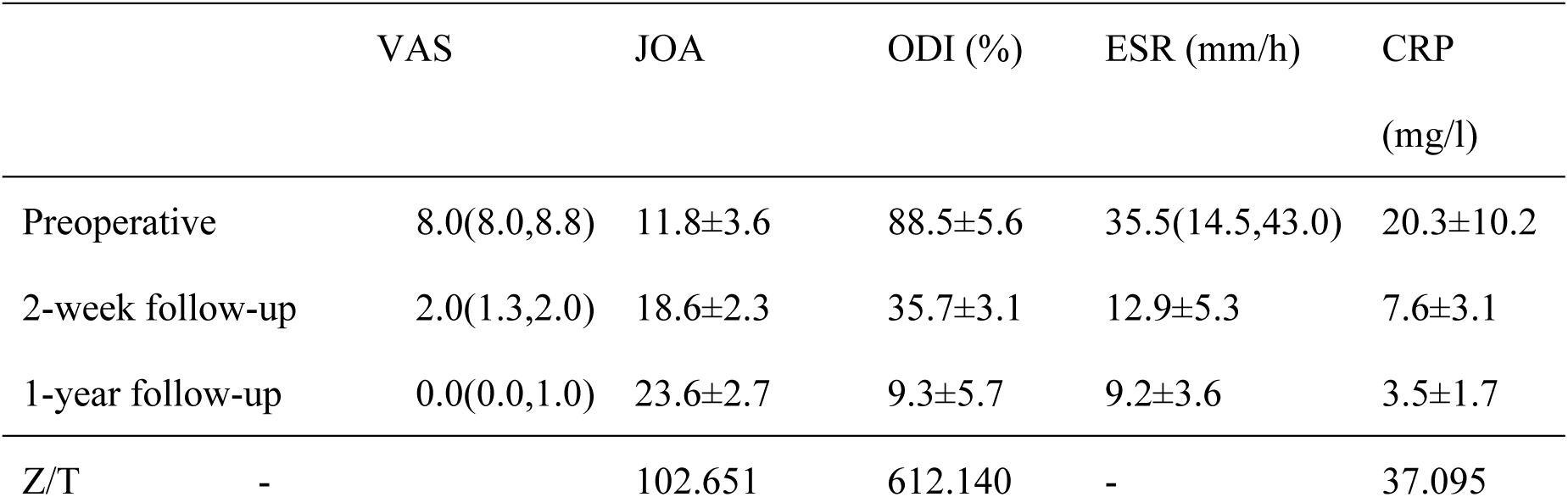
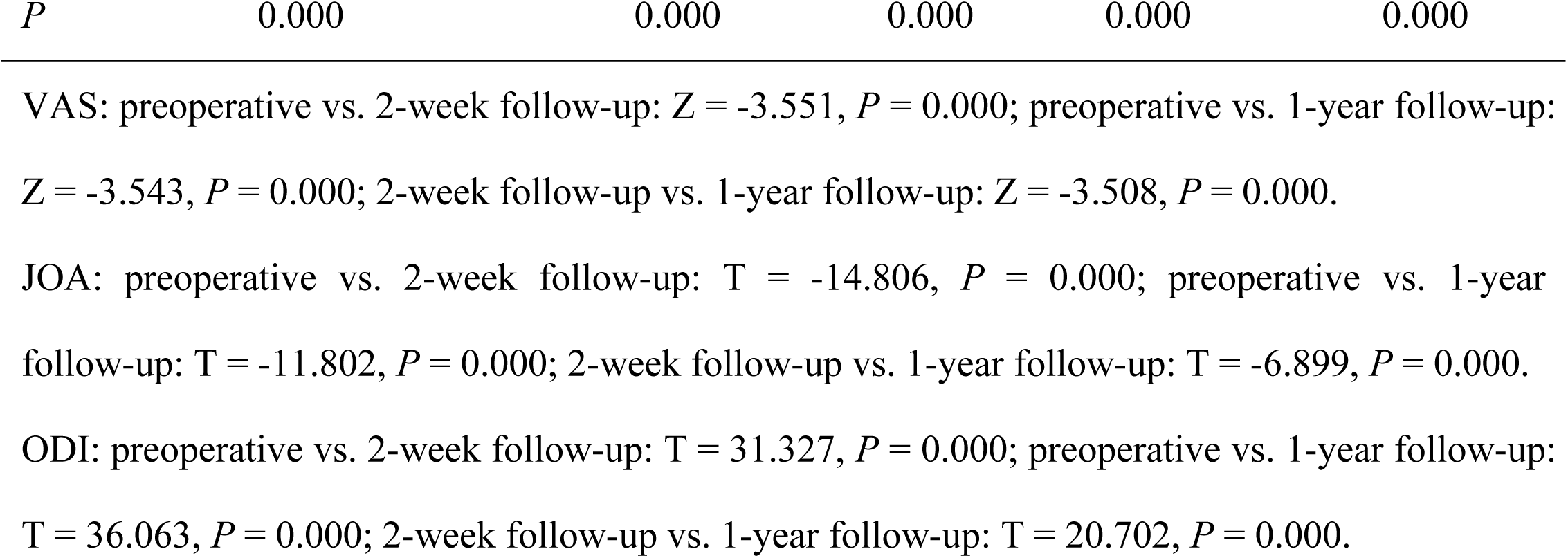
Comparison of preoperative and postoperative VAS, JOA, ODI, ESR and CRP.

|  | VAS | JOA | ODI (%) | ESR (mm/h) | CRP<br>(mg/l) |
| --- | --- | --- | --- | --- | --- |
| Preoperative | 8.0(8.0,8.8) | 11.8±3.6 | 88.5±5.6 | 35.5(14.5,43.0) | 20.3±10.2 |
| 2-week follow-up | 2.0(1.3,2.0) | 18.6±2.3 | 35.7±3.1 | 12.9±5.3 | 7.6±3.1 |
| 1-year follow-up | 0.0(0.0,1.0) | 23.6±2.7 | 9.3±5.7 | 9.2±3.6 | 3.5±1.7 |
| Z/T | - | 102.651 | 612.140 | - | 37.095 |
| *P* | 0.000 | 0.000 | 0.000 | 0.000 | 0.000 |

VAS: preoperative vs. 2-week follow-up: $Z = -3.551$ , $P = 0.000$ ; preoperative vs. 1-year follow-up: $Z = -3.543$ , $P = 0.000$ ; 2-week follow-up vs. 1-year follow-up: $Z = -3.508$ , $P = 0.000$ . JOA: preoperative vs. 2-week follow-up: $T = -14.806$ , $P = 0.000$ ; preoperative vs. 1-year follow-up: $T = -11.802$ , $P = 0.000$ ; 2-week follow-up vs. 1-year follow-up: $T = -6.899$ , $P = 0.000$ . ODI: preoperative vs. 2-week follow-up: $T = 31.327$ , $P = 0.000$ ; preoperative vs. 1-year follow-up: $T = 36.063$ , $P = 0.000$ ; 2-week follow-up vs. 1-year follow-up: $T = 20.702$ , $P = 0.000$ .

### Comparison of preoperative and postoperative ESR and CRP

ESR and CRP gradually decreased to normal in 16 patients. ESR decreased from 35.5 (14.5,43.0) mm/h preoperatively to (12.9 ± 5.3) mm/h and (9.2 ± 3.6) mm/h at 2 weeks and 1 year after the operation, respectively. CRP decreased from (20.3 ± 10.2) mg/l preoperatively to (7.6 ± 3.1) and (3.5 ± 1.7) mg/l at 2 weeks and 1 year after the operation, respectively. ESR and CRP at 2 weeks and 1 year after the operation were significantly different from those before the operation (*P* = 0.000) (**Table 2**). Moreover, 1 year after the operation, ESR and CRP were significantly different from those 2 weeks (*P* = 0.000). (**Table 2**)

### Radiological evaluation

The average spondylolisthesis reduction rate 2 weeks after operation was (91.2 ± 6.7) %, and the median reduction loss rate 1 year after operation was 8.0 (5.0,9.8) %. At the last follow-up, all patients achieved interbody fusion without loosening or fracture of instrumentation, and no patient had recurrence (**Fig. 2**).

## Discussion

The pathological changes of brucella spondylitis are mainly infectious vertebrates and discitis, with vertebral destruction, spinal instability, and spinal cord or nerve compression [3-5,15]. In recent years, with further research on spinal biomechanics, researchers believed that surgery should be carried out to remove the lesions and reconstruct spinal stability to relieve pain and promote the recovery of nerve function when antibiotic treatment is not enough for curing [7,16,17]. Tebet [18] classified the causes of spondylolisthesis as isthmic, degenerative, and traumatic, especially isthmus and degeneration. The incidence of lumbar spondylolisthesis in the general population is about 2% ∼ 6%. However, few studies on the correlation between brucella spondylitis and lumbar spondylolisthesis have been done. There is also a lack of statistical data on the incidence of lumbar spondylolisthesis in patients with brucella spondylitis. As we all know, the spine’s stability depends on the joint maintenance of the complete vertebral body, intervertebral disc, pedicle, ligament, and surrounding tissue. We believe that brucella spondylitis mainly invades the adjacent vertebral upper and lower endplates and intervertebral discs, which easily destroy the spine’s stability, aggravate the degeneration, and then cause spondylolisthesis. In this study, 10 of the 16 patients (62.6%) were combined with isthmus, with a high proportion, which is one of the main factors of lumbar spondylolisthesis.

Clinically, various surgical methods for brucella spondylitis include anterior, posterior, or anterior and posterior debridement for bone graft fusion, and minimally invasive surgical methods, which emerged in recent years [7,16,17,19,20]. The traditional surgical methods of brucella spondylitis are anterior and posterior combined debridement, bone graft fusion, and instrumentation. This method has obvious disadvantages, including long operation time, changing body position during operation, and too complex operation [21,22]. Therefore, more surgeons have chosen posterior debridement, bone graft fusion, and instrumentation in recent years. This operation avoids the shortcomings of traditional operations and can significantly reduce the probability of recurrence [16,17,19]. The surgical methods of lumbar spondylolisthesis are also diverse. The most classic surgical methods are posterior interbody fusion. Transforaminal and extreme lateral approaches gradually developed, including the oblique lateral approach, which has sprung up in recent years, are posterior. The surgical core is reduction, decompression, fixation, and fusion [23,24]. So, we adopted a posterior approach in this study.

The spine’s stability needs a typical spinal sequence to maintain, but there are still disputes about whether the spondylolisthesis needs reduction and the degree of reduction. Poussa et al. [25] showed that in situ fusion of spondylolisthesis vertebral body could also achieve satisfactory clinical results, while spondylolisthesis reduction might bring discomfort for patients. On the other hand, Dewald et al. [26] reported that complete reduction would lead to excessive nerve root traction. Therefore, a partial reduction of the spondylolisthesis vertebral body should be recommended. More researchers [27,28] suggested reducing the spondylolisthesis vertebral body to the greatest extent, which will help restore the typical spinal sequence and promote bone graft fusion. As the postoperative recovery of brucella spondylitis needs stability, we reduced the spondylolisthesis vertebral body during the operation as much as possible in this study.

However, the reduction is temporary, and bone graft fusion is the ultimate goal. The use of internal fixation instruments could maintain the reduction of the spondylolisthesis vertebral body, and bone graft fusion would help the stability of the spine [29]. Dantas et al. [30] found that posterolateral bone graft was not on the load-bearing axis, which was prone to the non-fusion of bone graft or formation of the pseudo joint. Through a follow-up study, Miyashita et al. [31] found that pedicle screws and interbody fusion cage could rebuild spinal stability, effectively maintain the height of intervertebral space, reduce screw pressure, and reduce the incidence of fracture and loosening of pedicle screws. Compared with simple debridement and bone graft fusion, instrumentation could make the local stability conducive to bone graft fusion and promote lesions cure, with fewer postoperative complications and a low recurrence rate [7,17,32]. Bone graft fusion is the basis of spinal stability. Therefore, internal fixation instruments are conducive to bone graft fusion. Said et al. [33] researched that the patients with lumbar spondylolisthesis combined with an isthmus, posterior interbody fusion might restore spinal stability, improve bone graft fusion, and have an excellent long-term clinical effect. In this study, 62.6% of patients were complicated with an isthmus. Therefore, we adopted polyetheretherketone (PEEK) cages, interbody fusion, and pedicle screw fixation.

The postoperative follow-up showed that the average spondylolisthesis reduction rate in 16 patients was (91.2 ± 6.7) % two weeks after the operation. The median reduction loss rate was 8.0 (5.0,9.8) % one year after the operation, indicating the beneficial influence of the surgery. At the last follow-up, all patients had an interbody fusion, no loosening, fractures of instrumentation were found, and no patient had a recurrence, which means satisfactory clinical outcomes. During the follow-up, the clinical symptoms of 16 patients gradually improved, and the inflammatory indexes were back to normal. In addition, we found that the VAS score, JOA score, ODI index, ESR, and CRP at two-week and one-year follow-up were significantly different from those before the operation, which showed a marked improvement in the patient’s condition. Moreover, VAS score, JOA score, ODI index, ESR, and CRP at one-year follow-up were significantly better than those at two-week follow.

Our study had limitations: the number of patients was small, the evaluation indicators were insufficient, and the possibility of bias hampered the retrospective studies. Therefore, further observation, research, and clinical evaluation of cohort studies are needed.

In conclusion, a single-stage posterior approach could achieve a beneficial outcome for patients with lumbar brucella spondylitis combined with spondylolisthesis. Posterior lumbar debridement, reduction, interbody fusion, and instrumentation could reconstruct the spine’s stability, relieve pain and promote the recovery of nerve function.

## Data Availability

All relevant data are within the manuscript and its Supporting Information files.

N

## Acknowledgements

The authors thank the participants for making this study possible and the Department of Pathology for technical support. This research was funded by the science foundation of Beijing Ditan Hospital Capital Medical University (No.DTQL201803).

## Ethics approval and consent to participate

Ethical approval from the Ethics Committee of Beijing Ditan Hospital, Capital Medical University was obtained for this study. Each author certifies that all investigations were conducted in conformity with ethical principles.

## Author contributions

YZ and CSZ conceived the manuscript, participated in the collection of clinical data and manuscript writing; YZ and JMC participated in the pathological examination, YZ and QZ participated in the manuscript revision. All authors read and approved the final manuscript.

